# Individuals with intellectual disability have an increased burden of *de novo* variants in *cis*-regulatory elements of glutamatergic neurons

**DOI:** 10.1101/2025.09.23.25336266

**Authors:** Eric A. Sosa, Dónal O’Shea, Samuel Rosean, Caroline Yoon, Jacob Stauber, Pablo E. Castillo, Cathal Seoighe, Melissa J. Fazzari, John M. Greally

## Abstract

To understand the role of non-coding sequence variation in neurodevelopmental disorders, we studied 709,222 *de novo* variants (DNVs) using the genomes of 20,366 individuals. We found enrichment of DNVs at *cis*-regulatory elements (CREs) of all somatic cell types, attributable to the deamination of 5-methylcytosine at CG dinucleotide-dense CREs. In neurodivergent individuals with or without intellectual disability (ID), the DNVs were enriched at genes implicated in autism/ID at promoter-proximal CREs of brain cell types specifically. As the properties of the DNVs were not sufficiently distinctive to explain the phenotypic associations, we performed a multivariate category-wide association study (CWAS) that revealed enrichment of DNVs in CREs of glutamatergic neurons in individuals with ID, with a random forest analysis demonstrating an increased burden of these events in these individuals with ID. Our findings are consistent with a model of non-coding variants contributing to the oligogenic heritability of neurodevelopmental disorders.

## MAIN

The current understanding of the heritability of autism^1^ and ID^2^ is one of multiple genetic events combining to cause the phenotype.^3,4^ These oligogenic models are based on damaging variants found in the interpretable protein-coding minority of the human genome. How variants in the non-coding remainder of the genome contribute to neurodevelopmental disorders is largely unknown, although some studies have implicated non-coding *de novo* variants (DNVs) in both autism^5^ and ID.^6^ With increasingly large cohorts of individuals studied using whole genome sequencing (WGS),^7^ among them cohorts with neurodevelopmental phenotypes,^8,9^ combined with improved insights into the regulatory landscapes of multiple cell types,^10^ including those of the human brain,^11^ we have the opportunity to improve our understanding of the role of variants in non-coding regulatory elements in neurodevelopmental phenotypes.

We studied 20,366 individuals from three cohorts: MSSNG,^9^ the Simons Simplex Collection (SSC)^8^, and TOPMed.^7^ We focused on the 709,222 *de novo* single nucleotide variants (SNVs) and small insertions or deletions (indels) identified in these studies. We used a filtering strategy to limit technical artifacts and excluded neurodivergent individuals with a recognized genetic cause for their phenotype (‘solved’ cases), leaving us with 597,439 unique DNVs from 7,535 unsolved individuals for further analysis. Of these individuals, 5,415 were categorized as having the trait of autism, of whom a subset of 916 were further categorized as intellectually disabled (annotated with a full-scale IQ ≤70), with 2,337 described as having typical IQ (>70).

Our first question was whether the DNVs in individuals with autism were enriched in the regulatory regions of specific cell types, where they could mediate functional effects on transcriptional regulation. For this, we used our Regulatory Landscape Enrichment Analysis (RLEA) approach^12^ to test for non-random over-representation of DNVs in open chromatin regions of 44 cell types. In contrast to our prior application of RLEA to genome-wide association study variants,^12^ every cell type tested revealed significant enrichment for DNVs in their open chromatin regions in individuals with autism and in neurotypical controls (**Supplementary Fig. 1**).

This raised the question of how an event likely to occur during gametogenesis could select loci destined to have regulatory in derived somatic cells. The role of base composition became apparent with the mutational profiling of the subset of 527,717 single nucleotide substitutions using SigProfilerMatrixGenerator,^13^ which revealed enrichment for the SBS1 and SBS5 signatures (**Fig. 1a**), a finding previously reported for human DNVs.^14–16^ Both are described as “clock-like” signatures of sequence variation associated with age. SBS1 represents C>T transitions due to the deamination of 5-methylcytosine at CG dinucleotides, while SBS5 appears to be due to errors in DNA repair.^16,17^

**Figure 1:**
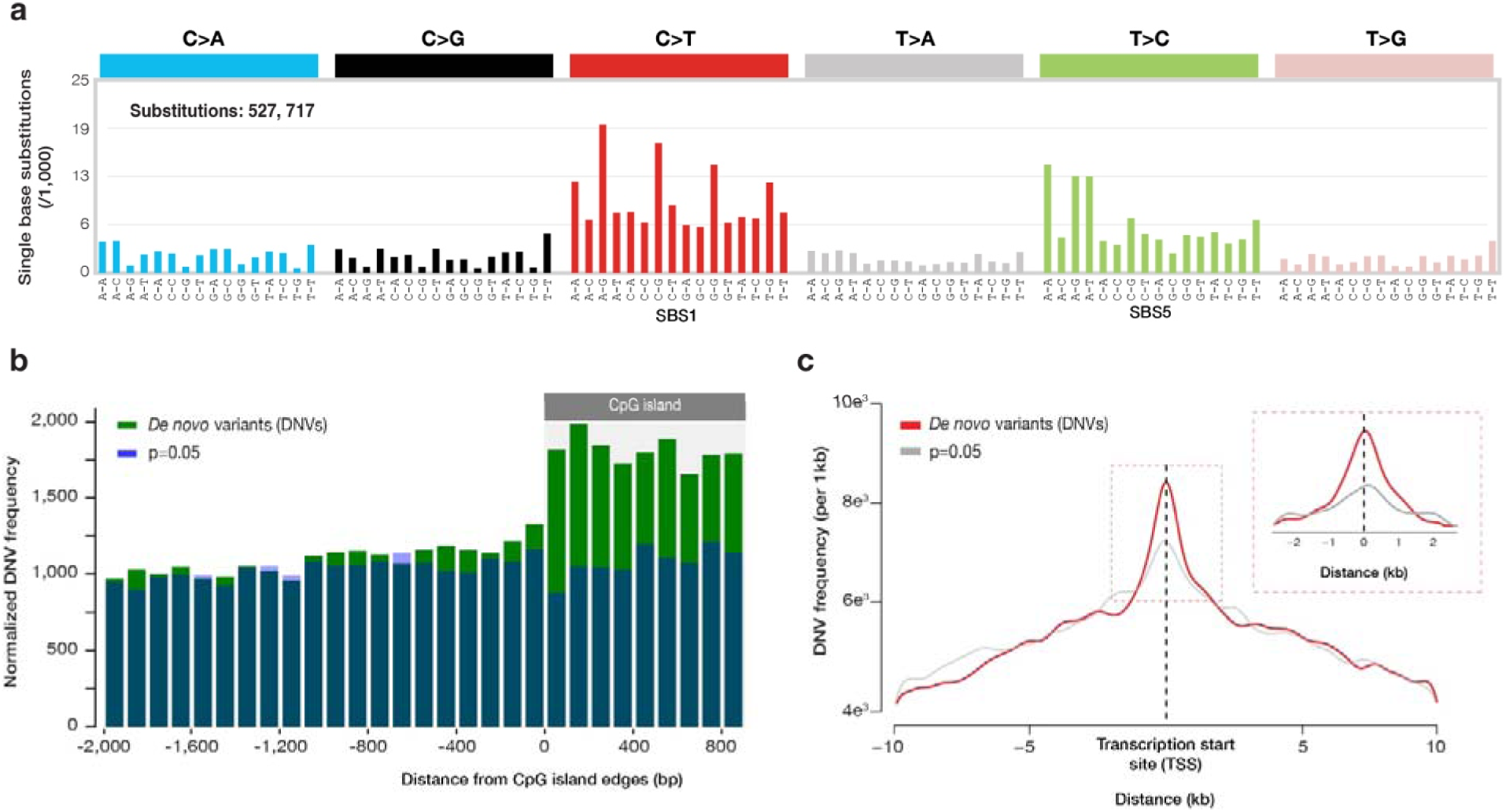
D**N**V **enrichment within CpG islands and at the TSS is driven by C>T transitions at CG dinucleotide-rich loci. (a)** Mutational signature analysis of 527,717 *de novo* single base substitutions revealed a pattern of enrichment for C>T transitions associated with the SBS1 (red) and the T>C transitions of SBS5 (green) signatures. **(b)** Even after normalising for the increased density of CG dinucleotides, DNVs are significantly enriched in CpG islands (green bars). The frequency of observed DNVs (green) is significantly greater than that of the p=0.05 threshold (blue) in CpG islands. **(c)** DNVs are also significantly enriched in the ±2 kb region flanking annotated TSSs compared with the p=0.05 threshold (gray line).

The enrichment for events at CG dinucleotides suggested a mechanism for the targeting during gametogenesis of *cis*-regulatory elements (CREs) active in derived somatic cells, as CREs tend to have increased CG dinucleotide content.^18^ When we tested for enrichment within CpG islands, we found an increased number of DNVs, even after normalizing for the increased CG dinucleotide content of CpG islands (**Fig. 1b**), and enrichment in the ∼2 kb flanking transcriptional start sites (TSSs) (**Fig. 1c**). Human DNVs therefore disproportionately target CREs through deamination of 5-methylcytosines, leading to enrichment at these CG dinucleotide-enriched regions.

To refine the analysis further, we asked whether the DNVs in the individuals with autism were occurring within CREs near the subset of genes that have previously been associated with autism through damage to their coding sequences. We started with the 162 autism genes defined by the SPARK autism research study^19^ and tested whether DNVs were occurring in regulatory loci of any of the 44 cell types within 10 kb of the TSSs of these 162 genes (promoter-proximal CREs (ppCREs)). This revealed selective enrichment in individuals with autism for regulatory loci in specific brain cell types, both neuronal and glial, as well as in tissue samples representing mixtures of cells from several brain regions (**Fig. 2a**). We focused on the five individual brain cell types (including a combined microglia/astrocyte dataset) to test whether the finding for the 162 SPARK genes was consistent for 14 other autism gene lists (**Supplementary Table 1**). This showed that DNVs in individuals with autism consistently target ppCREs in glutamatergic neurons for all the autism gene lists, and that the brain cell types tested have increased targeting of CREs by DNVs for most of the gene lists (**Fig. 2b**).

**Figure 2:**
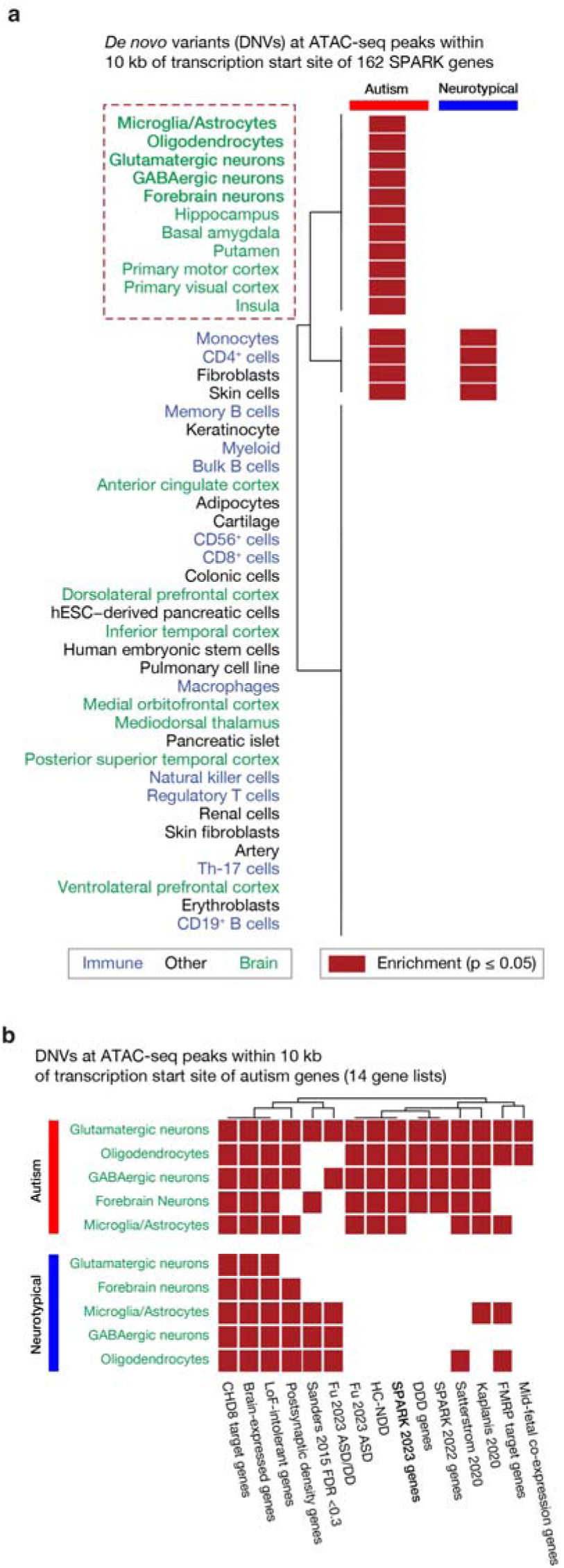
D**N**Vs **in autism preferentially occur in *cis*-regulatory loci of brain cells near known autism genes. (a)** A study of the DNVs at ppCREs near SPARK genes across 44 cell types identified a subset of cell types to be enriched in individuals with autism. The heatmap demonstrates the 11 brain cell types and brain regions that significantly distinguishes the autism and neurotypical groups (highlighted by red dashed line box). **(b)** Testing the five brain cell types from the analysis in (a) in a broader set of 15 autism gene lists reveals a similar enrichment of DNVs in ppCREs for most of the gene lists.

With evidence that DNVs in individuals with autism are targeting ppCREs of genes previously associated with autism, we then asked whether the DNVs in individuals with autism were qualitatively different from those in neurotypical individuals and whether, within the autism group, the DNVs in the subset of individuals with low IQ were also distinctive. We focused on DNVs in ppCREs in the brain cell types highlighted in **Fig. 2b**. We compared these DNVs between the individuals with autism and neurotypical individuals, and the subgroups with low (≤70) and typical (>70) IQ within the autism group. We studied evolutionary conservation, distance to the nearest transcription start site or the summit/center of the peak of open chromatin, and an index we created that quantifies how shared each regulatory element is across cell/tissue types. While there were some differences observed between groups (**Supplementary Figs. 2-3**), the innate properties of the DNVs did not obviously distinguish the different phenotypic associations.

As the individual properties of the DNVs were not providing clear reasons for the phenotypic differences, we implemented a more comprehensive multivariate analysis to study the associations between the DNVs and both the neurodivergent and ID phenotypes. We adopted the category-wide association study (CWAS) approach that was previously developed to study the role of non-coding DNVs in the SSC cohort.^5,20^ We modified some of the published peak calling assumptions to update the functional annotation categories of the prior studies, and then used the SSC cohort alone to replicate successfully the previously described strong signals for *de novo* protein-truncating and missense variants in genes implicated in autism in this group^20^ (**Supplementary Fig. 4**). We then replaced the functional categories that were uninformative in this SSC replication analysis with the ATAC-seq data that were informative in our RLEA analysis and added two further descriptive categories (proximity to the TSS and AlphaMissense^21^ prediction of damage to the coding sequence, **Supplementary Fig. 5**). We then performed the resulting CWAS on the filtered 597,439 unique DNVs from the 7,535 individuals from the combined SSC, MSSNG and TOPMed cohorts. A comparison between DNVs in ppCREs from individuals with autism and those of neurotypical individuals was consistent with the prior study from An *et al.,*^5^ who found associations for non-coding variants in the autism group to be substantially less significant than for *de novo* protein-coding variants. The autism group had increased DNV accumulation compared with the neurotypical group at DNase hypersensitive sites identified by the ENCODE project^22^ near genes from the Brain-expressed list at *p* values <0.0001 (**Supplementary Fig. 6**, **Supplementary Table 2**), exceeding the *p* values of the pathogenic, causative *de novo* missense variants shown in **Supplementary Fig. 4**.. In contrast, when we compared the smaller subset of individuals with low (≤70) and typical (>70) IQ within the autism cohort, three associations emerged as comparably positive. DNVs in individuals with low IQ were enriched in ppCREs of several types of glutamatergic neurons (plus hippocampal tissue), linked to genes in three of the autism gene lists (**Fig. 3a**, **Supplementary Table 3**).

**Figure 3:**
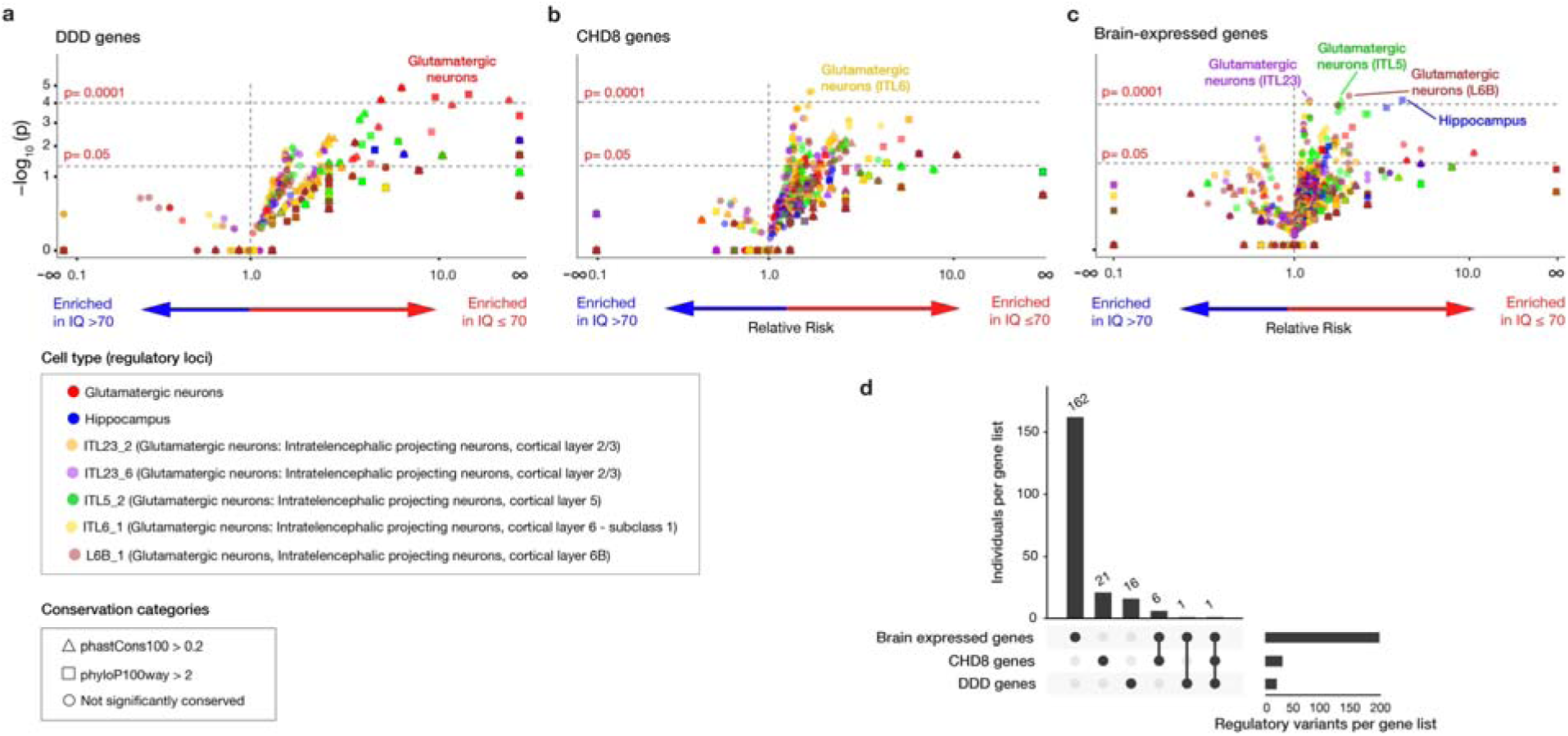
CWAS reveals DNV enrichment at CREs of glutamatergic neurons near developmental delay genes and implicates mid-fetal developmental timing effects in ID. **(a)** Glutamatergic DNVs in ppCREs (red points) near known developmental delay genes (DDD) are significantly enriched in the low (≤70) IQ group, a finding consistent with the results of pseudobulked ATAC-seq from single nuclear studies of human brain for different types of glutamatergic neurons proximal to the CHD8 and Brain-expressed gene lists also implicated in autism. Linking these variants back to the individuals studied reveals the categories of panel (c) to be the most represented among the 207 study participants with these variants and low IQ.

When linked to the individuals in whom the DNVs were found, the largest group of individuals was those with DNVs proximal to Brain-expressed genes (**Fig. 3d**).

These studies of DNVs aggregated from thousands of individuals in each tested group revealed properties of the DNVs themselves, but without insights into how they may mediate an individual’s phenotype. We therefore linked DNVs within CREs and their descriptive properties to each research participant, along with other descriptors of the individual (**Supplementary Tables 4-5**). A random forest model testing the differences between the low and typical IQ groups showed the strongest effects to be mediated by the relative contribution of the total number of DNVs in an individual within ppCREs of glutamatergic and GABAergic neurons (**Fig. 4a**). We explored this further, showing that the proportion of individuals in the lower (≤70) IQ group correlates with an increasing number of DNVs in ppCREs of glutamatergic neurons in that individual’s genome (**Fig. 4b**). This is consistent with results of a prior study of 242 individuals with autism and SSC probands^23^ which showed a similar association between the burden of DNVs at loci with regulatory properties and low IQ.

**Figure 4.**
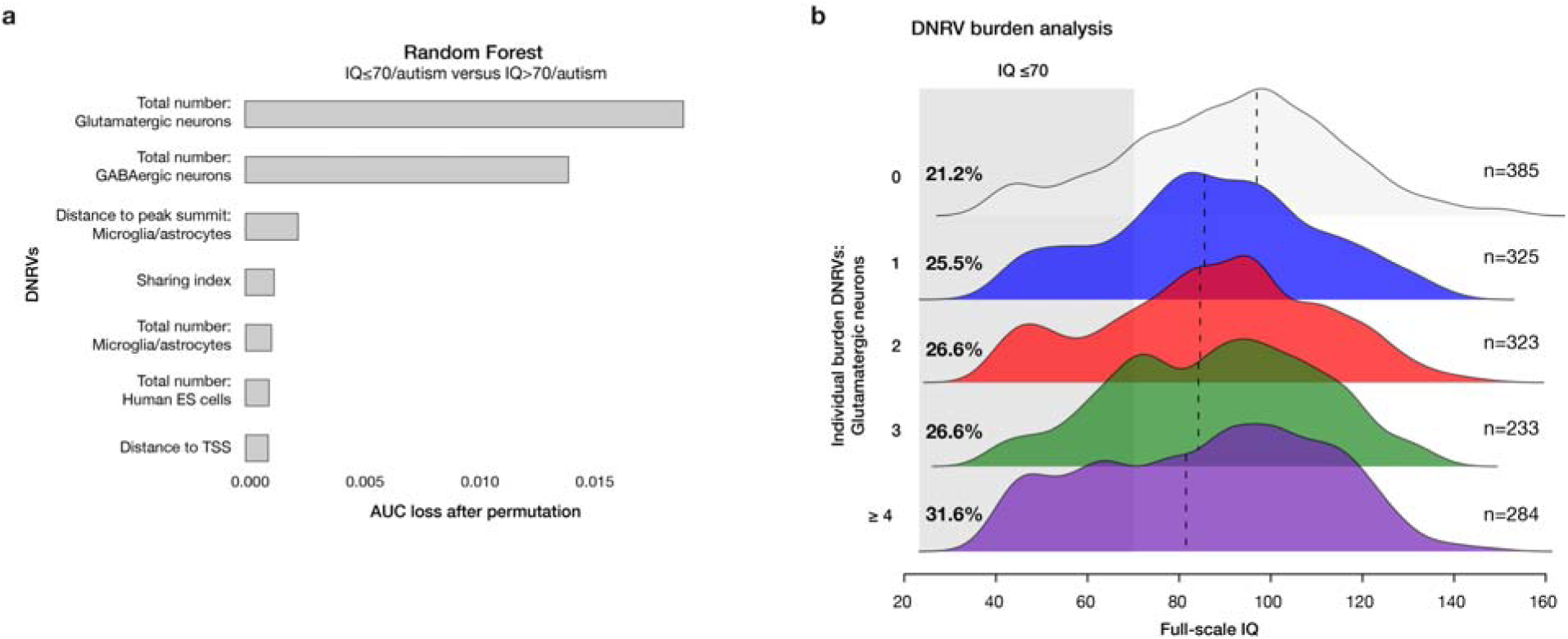
Individual-focused modelling shows the effect of DNV burden in CREs of glutamatergic neurons on intellectual impairment. **(a)** A random forest permutation-based approach was performed to reveal functional categories distinguishing individuals with low and high IQ. The amount of change of the area under the curve (AUC) was assessed for each functional category after permutation. The strongest effects were found for the total number of DNVs in ppCREs of glutamatergic neurons, followed by GABAergic neurons. **(b)** The burden of DNVs in ppCREs of glutamatergic neurons is negatively associated with full-scale IQ. The average IQ score of each group of individuals with 0, 1, 2, 3, and ≥ 4 DNVs in ppCREs of glutamatergic neurons is shown by the vertical dashed line. An increase in the proportion of individuals with intellectual impairment (bolded percentages) is seen as the number of DNVs in ppCREs of glutamatergic neurons increases.

When the individual-level clinical relevance of non-coding DNVs was tested in a prior deep learning study, it was estimated that an additional 4.3% of the individuals with autism in the SSC cohort had their phenotype explained by the *de novo* non-coding mutations predicted by the deep learning model.^24^ Using the same assumption of a causal relationship for non-coding DNVs, focusing on the DNVs highlighted as significant in the CWAS analysis of **Fig. 3**, we identified the individuals in whom the variants occurred (**Fig. 3d**) and quantified their proportion within the cohort studied in this project. These 207 unsolved individuals comprise an additional 3.8% of the 5,415 individuals with autism, comparable with the additional 4.3% described in the deep learning model study.^24^ However, for the 916 individual with IQ ≤70, the 207 individuals with DNVs defined as significant in the CWAS analysis, these represent an additional 22.6% (**Fig. 5**), substantially exceeding the separate 15% (138/916) subset of individuals with ID who were categorized as solved.

**Figure 5.**
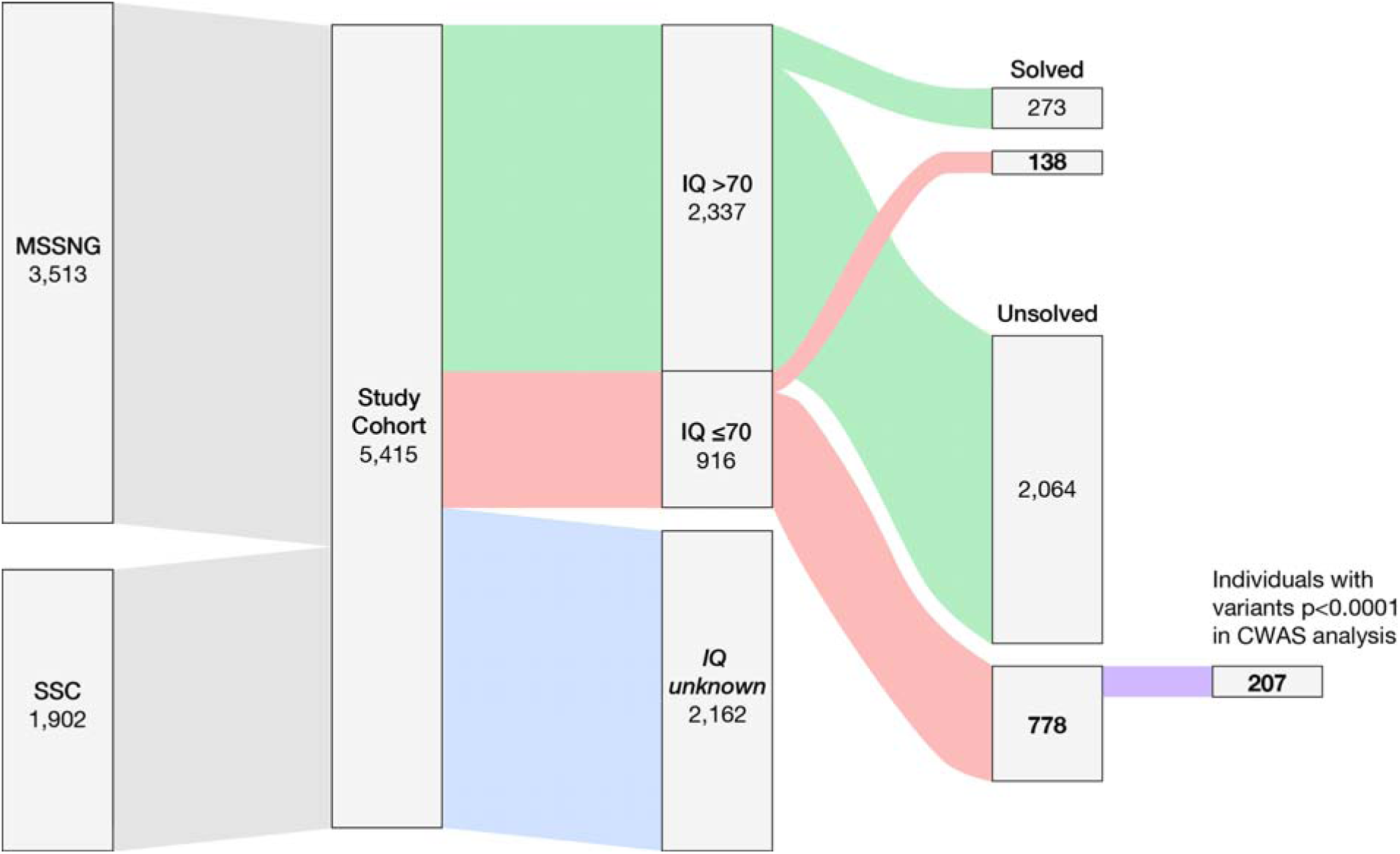
Using DNV information in ppCREs may increase the diagnostic rate in intellectual disability. We show the composition of the cohorts, breaking down those with IQ measurements, and those in the intellectually disabled (IQ≤70) and typical IQ (>70). The solved subgroups are defined by pathogenic and causative genetic variants explaining the autism trait, of whom a subset have intellectual disability. Of the 778 unsolved individuals with IQ≤70, 114 have DNVs in ppCREs of glutamatergic neurons near DDD genes^25^ or in ppCREs of mid-fetal brain development near genes co-expressed at that stage.^26^. Of the 916 individuals with IQ≤70, these 114 individuals represent an additional 12.4% on top of the 138 (15%) with diagnostic genomic changes (solved).

This study, which we believe to be the largest to date testing the role of DNVs in autism and ID, reveals how DNVs non-randomly target CREs in the human genome, mediated by variants at CG dinucleotides. We show that in a cohort selected for having the trait of autism, non-coding DNVs are enriched in regulatory regions of brain cells and tissues at genes already implicated in causing this phenotype through coding sequence effects. Overall, the properties of the DNVs did not discriminate the autism or low IQ groups but appeared to have enough distinctive patterns to warrant a multivariate analysis to understand how they might contribute combinatorially to the autism and ID phenotypes. We chose the CWAS approach that was previously developed to study the role of DNVs in autism,^20^ updated to use new functional genomic data defining the regulatory regions of brain cell types, revealing the strongest effects for DNVs in regulatory loci of glutamatergic neurons when we focused on individuals with ID. To understand further the influences of the DNVs on phenotypes, we collected information about each individual study participant and found an effect of DNV burden, those individuals with a larger number of DNVs at CREs of glutamatergic neurons having a higher chance of intellectual disability.

Our focus on the SNV and indel subset of DNVs was designed to understand the influence of smaller variants that are more readily linked to individual ppCREs and genes. This focus revealed an unexpected enrichment of these smaller DNVs at regulatory loci in all cell types studied. The mutational signatures of these DNVs included a strong over-representation of the cytosine to thymine transition pattern in the context of CG dinucleotides, attributable to the deamination of 5-methylcytosine.^27^ This effect of base composition explains why events during gametogenesis occur at loci that will become regulatory elements later in development, without the need for a local chromatin or other transcriptional regulatory state to be distinctive during gametogenesis. The observation of Vierstra and colleagues that there is increased sequence polymorphism immediately at transcription factor binding sites in somatic cells,^28^ suggests support for the proposed model of competition between transcription factors and DNA damage repair enzymes.^29,30^ For this model to be related to DNV targeting of CREs of somatic cells, the transcription factor binding found in somatic cells would also have to occur during gametogenesis, making this model less likely than one simply based on CG dinucleotide content.

Our finding of DNVs occurring at increased frequency at *cis*-regulatory elements at genes implicated in a disease phenotype expands on prior comparable studies in patients with cardiomyopathies,^31^ congenital heart disease,^32^ systemic lupus erythematosus,^33^ and autism.^5,23^ The DeepSEA deep learning-based model applied to DNVs in the SSC cohort found an association between DNVs in regulatory regions and lower IQ.^24^ In a study of rare recessive variants, enrichment at non-coding regulatory regions was also found in individuals with autism.^34^ Our findings are consistent with these multiple prior studies while allowing a focus on glutamatergic neurons as a major mediating cell type, with mid-fetal development as the critical time for DNVs to have deleterious consequences, and a burden of DNV load at regulatory loci associated with the low IQ phenotype.

This study implicates glutamatergic neurons in the pathophysiology of ID. Glutamatergic pathways have been strongly associated with autism and ID,^35^ while developmental studies in organoid models indicate that some of the effects may not be intrinsic to the glutamatergic neurons but instead related to the imbalance of differentiation of excitatory GABAergic with inhibitory glutamatergic neuronal differentiation.^36^ The indication of mid-fetal gestation as the critical timing for DNV effects is consistent with the expression patterns of genes implicated in autism, in particular those expressed in layer 5/6 cortical projection neurons.^26^ A separate study of copy number variation of the 16p11.2 region, responsible for neurodevelopmental phenotypes, found effects in late mid-fetal brain development involving glutamatergic neuron markers in cortical layers 5 and 6.^37^ Our results show enrichment for DNVs in regulatory regions of glutamatergic neurons, and for DNVs in regulatory regions of mid-fetal brain development, supporting a potential role for non-coding variants in the disruption of development of these cortical layers and a resulting common outcome of neurodevelopmental disorders.

The results of this study are consistent with a genomic architecture of ID involving multiple events co-occurring to confer disease risk. Our model of an increased burden of DNV in regulatory regions of glutamatergic neurons associated with a higher risk of ID is consistent with studies focused on burdens of damaging coding sequence variants, together supporting an oligogenic model for ID.^38–40^ It is likely that coding and non-coding variants combine to confer risk, potentially allowing future burden analyses to combine both types of variants in diagnostic models of ID. A goal will be to move beyond DNVs to study other rare variants in regulatory regions, for which predictive models such as DeepSEA^41^ will be valuable. This study supports the need to use large cohorts with genome sequencing and the ability to call DNVs for discovery, which will be aided in future studies by the greater numbers of DNVs identifiable using new long-read sequencing technologies.^42^ The combination of new sequencing technologies, diagnostic sequencing of coding and non-coding DNA, the development of predictive models, and the insights into the contribution of non-coding variation to these models should combine to improve significantly our ability to deliver diagnostic insights for individuals with ID and their families.

## ONLINE METHODS

### Whole genome sequencing data

To understand the contributions of genetic variants to autism and intellectual disability (ID), we analyzed 360,791 DNVs from 11,312 individuals from the MSSNG cohort.^9^ This dataset included 5,100 individuals with autism (4,073 males, 1,027 females) and 6,081 neurotypical individuals (3,035 males, 3,046 females). 1,419 of the 5,100 individuals with autism were from multiplex families (two or more siblings with autism) while the remaining were from simplex families (single child with autism). Variant calls, phenotype data, annotations, and metadata were downloaded from the MSSNG portal (https://research.mss.ng/). To enhance the statistical power of our analyses, we analyzed an additional 255,106 DNVs from 1,902 quartet families – composed of two parents, a neurotypical sibling, and a child with autism – from the Simons Simplex Collection (SSC).^8^ The SSC cohort consists of WGS from 7,608 genomes (3,804 parental genomes, 1,902 neurotypical siblings, and 1,902 individuals with autism) that are available via the Simons Foundation Autism Research Initiative (SFARI, https://www.sfari.org/resource/sfari-base/).

As an apparently neurotypical control dataset, we used whole genome sequencing (WGS) from the Trans-Omics for Precision Medicine (TOPMed) consortium, analyzing 93,325 DNVs from 1,446 individuals across five cohorts: the Framingham Heart Study (FHS), Genetic Epidemiology of Asthma in Costa Rica (CRA), Cleveland Family Study (CFS), Barbados Asthma Genetic Study (BAGS), and the Genetics of Cardiometabolic Health in the Amish (AMISH).^43^

### Variant and study participant filtering

Whole blood-derived DNA sequences were used for analyses. Due to their propensity to acquire variants *in vitro*, sequencing data derived from cell lines were excluded.^44^ Individuals with ≥150 DNVs were removed to limit false-positive DNVs due to sequencing errors or other factors. To assess the contribution of variants in the non-coding genome, we filtered individuals that were negative for known deleterious variants (large CNVs and *de novo* loss of function events) with the objective of studying *de novo* variants in individuals with autism for whom no genetic cause was determined. ‘Solved’ individuals (those with a known cause for their autism/ID) were removed and analyzed separately, while those in whom the autism diagnosis was unclear were excluded. Sequencing reads from the MSSNG and SSC cohorts were pre-aligned to the GRCh38 reference genome by the respective groups prior to our analyses, using comparable DNV calling approaches.^8,9^

### *De novo* variant mutational signature analysis

Mutational signatures for the 527,717 *de novo* single base substitutions from the MSSNG and SSC cohorts were tested using SigProfilerMatrixGenerator.^13^ Mutational patterns of single base substitutions were visualized using the plotSBS function in a Python environment. The single base substitution-96 (SBS-96) numerical plot graphed the trinucleotide contexts of each variant. The x-axis shows the six categories that represent the different single base substitutions (C>A, C>G, C>T, T>A, T>C, and T>G), while the y-axis represents the number of each mutation per 1,000 bases. Duplicates variants were filtered prior to analysis. Visualization of the generated matrices demonstrated enrichment for both SBS1 and SBS5 mutational signatures. The GRCh38 genome was downloaded using the BSgenome.Hsapiens.UCSC.hg38 R package.

### DNV enrichment at CpG islands

CpG island locations were downloaded online using the UCSC Table Browser data retrieval tool (cpgIslandExt table). The mean CpG island size was 780 bp with 95% of CpG islands less than 1,941 bp in size. A total of 597,439 DNVs were found to overlap 27,494 unique CpG islands using GenomicRanges.^45^ To test the frequency of overlap of DNVs with CpG islands and their flanking regions, upstream and downstream distances were combined to create a single plot with 100 bp bins. The distances of DNVs to the nearest CpG island edge were calculated and normalized based on the frequency of CpG islands represented within each 200 bp bin. For permutation analysis, DNVs were randomly redistributed across each chromosome 950 times using the permutation and random functions from NumPy, generating a significance threshold of p=0.05.

### DNV enrichment at transcription start sites (TSS)

Genomic coordinates for 43,106 TSS from the NCBI Reference Sequence (RefSeq) database,^46^ were related to the locations of 597,439 DNVs, analyzing those located within 10 kb of a TSS. Permutation analysis was performed as above to generate a p=0.05 enrichment significance threshold.

### Regulatory Landscape Enrichment Analysis (RLEA)

We have previously described Regulatory Landscape Enrichment Analysis (RLEA), which allows testing for significant enrichment of a set of DNA sequence variants across the open chromatin loci of multiple cell types.^12^ We tested 44 cell types derived from the ATACdb database^10^ and from a published dataset of ATAC-seq from human brain samples.^11^ All ATACdb datasets were converted to the GRCh38.p13 genome build using the UCSC Genome Browser LiftOver function. The results for 597,439 DNVs were visualized using a heatmap, highlighting cell types with overlap remaining significant after Bonferroni correction for multiple testing (*p* ≤ 2.0 x 10^-5^).

### Autism gene enrichment analysis

DNVs from individuals with and without autism were examined for enrichment throughout the regulatory regions of 44 cell types located near 162 SPARK genes (from the March 7 2023 list at https://sparkforautism.org/portal/page/spark-gene-list/). DNVs within open chromatin regions and near a SPARK gene TSS (±10 kb) were analyzed by hypergeometric testing for comparison between both groups. Hypergeometric analysis was performed using the phyper function.^47^ Statistical significance for each cell type was visualized by a heatmap (*p* ≤ 0.05).

The cell types significantly enriched for regulatory variants near SPARK genes – microglia/astrocytes, oligodendrocytes, glutamatergic neurons, GABAergic neurons, and forebrain neurons – were then analyzed by hypergeometric testing across 14 additional autism gene lists. Eight of these autism gene lists were derived from Supplementary Table 6 in the prior study by Werling and colleagues.^20^ These included genes in autism risk (FDR ≤ 0.1),^48^ developmental delay,^25^ brain expression (BE),^49^ autism co-expression networks,^26^ post-synaptic density (PSD) (from the Genes2Cognition database),^50^ 792 of the 842 published FMRP target genes,^51^ and CHD8 targets,^52,53^ and loss-of-function intolerance (pLI) genes (score ≥ 0.9).^54^ Six gene sets were added to expand the list of autism-related genes. These included 1,586 high confidence neurodevelopmental disorder (HC-NDD) genes.^55^, 285 developmental disorder (DD)-associated genes^56^, 102 genes implicated as autism risk genes (FDR ≤ 0.1)^57^, 60 genes achieving exome-wide significance (*p* ≤ 2.5 × 10^−6^)^58^, 72 autism-associated genes (FDR ≤ 0.001)^19^, and 373 genes developmental delay (DD) genes (FDR ≤ 0.001) enriched in progenitor and immature neuronal cell transcriptomes.^19^ The genes composing each list and their sources are summarized in **Supplementary Table 1**. Statistical significance for regulatory variant overlap across each gene set was visualised with a heatmap (*p* ≤ 0.05).

### Testing the properties of DNVs in regulatory loci of glutamatergic neurons

DNVs within glutamatergic regulatory regions were analyzed for conservation status (PhyloP100way and PhastCons100way), cell type-specificity (regulatory sharing index) and distance to a TSS (within ±10 kb) or to an ATAC-seq peak summit. Variant conservation scores – phastCons100 (ranging from 0 to 1) and phyloP100-way (ranging from-20 to 30) – were downloaded from the UCSC Table Browser.^59,60^ Conserved loci were defined by a PhastCons score ≥0.2 and/or PhyloP score ≥ 2 consistent with criteria applied in previous CWAS analyses.^5^

A ‘regulatory sharing index’ was developed to quantify the cell type specificity of variants in regulatory regions. This index ranges from 0 to 1, with scores of 0 indicating the regulatory locus to be present in only a single cell type, scores of 1 indicating it to be shared across all cell types tested. A regulatory sharing index was developed for each variant using pyranges.

To characterize variant profiles across autism severity, we compared these qualitive markers between neurotypical and autistic individuals, and autism in the presence and absence of ID (IQ ≤ 70). The most recent Wechsler and Full-Scale intelligence quotient (IQ) scores were used for individuals in the MSSNG and SSC cohorts, respectively. Logistic regression models were used to test the association between DNVs and distance to the nearest TSS and conservation status. Two-sided p-values corresponding to Wald tests for model coefficients were presented. The distribution of conversation scores and TSS distances (within a ±10 kb window) for regulatory DNVs were visualized using a density histogram.

### Category-wide association studies (CWAS) framework for 5,415 families

A category-wide association study (CWAS) framework^5,20^ was applied to test the association of DNVs with autism across 260,820 unique combinations of categories. Variants were annotated against 68 categories divided among 25 functional groups, 19 gene sets, 12 GENCODEv43 annotations^61^ (using Ensemble variant effect predictor (VEP; version 109)),^62^ 6 conservation measures, 3 descriptive categories, and 3 variant types. The functional annotation group included cell-type regulatory regions and variant proximity to peak summit for 25 diverse cell types and structures. Gene list annotations included SPARK genes, brain-expressed genes,^49^ FMRP targets^51^ and CHD8 targets,^52,53^ post-synaptic density genes,^50^ midfetal co-expression networks,^26^ developmental delay genes,^25^ constrained genes (pLI ≥0.9),^54^ protein-coding genes, pseudogenes, and long non-coding RNAs. GENCODE annotations were used to survey coding (frameshift, in-frame, missense, silent, loss-of-function) and non-coding categories (intergenic, intronic, promotor, 3’ and 5’ untranslated regions, other non-coding exons, and non-canonical splice sites). DNV characterization consisted of alpha missense score (likely pathogenic: ≥ 0.5),^21^ pLI score,^54^ loss-of-function observed/expected upper-bound fraction (LOEUF) scores (gene haploinsufficiency: < 0.2),^63^ gnomAD v3.1.2 allele frequency (ultra-rare ≤ 0.0001, rare: ≤ 0.01),^64^ JASPAR transcription factor binding-site (TFBS) profile (score ≥ 400, p ≤ 1.0 x 10^-4^),^65^ DNV in CRE sharing index, variant size (SNV, indel), distance to TSS, and regulatory region variant burden. Variant conservation was evaluated using phyloP100way (≥ 2.0), PhastCons100way (≥0.2), zoonomia ultra-conserved elements (zooUCEs), unannotated intergenic constrained regions (UNICORNs), and runs of contiguous constraint (RoCCs) scores^66^ ZooUCEs were defined as regions (≥20 bp) where all nucleotides shared identical base composition and align across ≥235 species. Constrained bases (phyloP FDR ≤0.05) fewer than 5 bp apart in unannotated intergenic regions predominately within 500 kb of a protein-coding gene TSS were designated as UNICORNs. Genomic regions under high constraints – where contiguous bases have a phyloP score ≥2.2 (FDR ≤0.05) – were categorized as RoCCs. All surveyed CWAS categories are shown in **Supplementary Fig. 5.**

### CWAS Analyses and Visualization

CWAS analyses used the statistical approach described Werling and colleagues.^5,20^ A two-sided exact binomial test was conducted for each combination of categories to test whether the relative risk (RR), the number of variants in the category in cases divided by the number of variants in the category in all samples, significantly differed from the proportions observed without respect to any category. Results from the CWAS were visualized using volcano plots to assess the extent of variant enrichment within each category. Each point represents the RR (x-axis) and significance of enrichment (y-axis) in cases (right x-axis) or controls (left x-axis) of variants overlapping a unique combination of categories. Statistical evidence for enrichment was indicated on the y-axis as the-log_10_ p-value, with red lines marking the threshold for nominal significance (p ≤ 0.05, p ≤ 0.0001). Visualizations were generated using the ggplot2, plotly, and NatParksPalettes packages in RStudio.

### Random Forest: identification of important features

A random forest classification model was employed to identify features contributing to autism and intellectual disability (ID) co-occurrence. The ensemble was comprised of 2,000 decision trees, with the mtry hyperparameter (number of candidate features considered at each split) set to the default for classification, *i.e.,* the square root of the total number of input features. To address class imbalance, ID cases were upweighted at a 4:1 ratio relative to controls.

Prior to model fitting, patient-level summary features were derived from variant-level data. These included total DNV burden: (1) across the genome, (2) within glutamatergic open chromatin regions, and (3) within 10 kb of transcription start sites (TSS). Additional features aggregated across all glutamatergic-region variants included both indicators and counts of variants within in brain-expressed and developmental delay gene lists, variant functional class, conservation scores (PhastCons > 0.20; PhyloP100 > 2), and maximum observed conservation scores.

In total, 42 features were included in the random forest model using all available samples (n = 2,833; 2,073 controls and 760 ID cases). The DALEX R package^67^ was used to compute permutation-based variable importance in a model-agnostic framework. To estimate relative importance, features were ranked by the drop in AUC observed when randomly permuted; the top-ranking features were presented. As the main objective of this analysis was feature discovery and interpretation, the overall model AUC summarizing predictive performance was not reported.

### Burden analysis of DNVs in ppCREs of glutamatergic neurons

IQ score distribution was examined as a function of the burden of DNVs in ppCREs of glutamatergic neurons in individuals with autism. IQ scores were stratified into groups according to the number of regulatory DNVs per genome. Individuals with 0, 1, 2, 3, and ≥4 DNVs in ppCREs of glutamatergic neurons were represented by different distributions. Density data were derived using kernel density estimation and presented using a ridgeline plot, where full scale IQ and individual glutamatergic ppCRE DNV burden were plotted on the x and y axes, respectively. The percentages on the left indicate the proportion of the group with intellectual impairment (IQ ≤ 70). The sample size of each group is provided on the right side of each distribution.

## AUTHOR CONTRIBUTIONS

E.A.S. gathered genomic datasets, generated data, conducted variant characterization analyses, provided figure visualizations, developed supplementary files, and contributed substantially to manuscript preparation. D.O. performed the trinucleotide context, mutational signature, and preliminary CWAS analyses in addition to extensive software modelling. S.R. facilitated variant enrichment analyses (RLEA), CpG and TSS burden calculations, and contributed scripts for chromatin analyses. C.Y. performed the CpG island analysis, J.S. provided GRanges scripts for variant-chromatin overlapping and CpG burden normalization, P.E.C. guided our interpretation of brain cell findings, C.S. provided statistical stringency and guidance during modelling and variant prediction efforts, M.J.F. performed all machine learning analyses, suggested code modifications to increase analytical power, and validated all statistical testing. J.M.G. acquired the WGS data, designed the research, supervised the project’s progress, provided figure visualizations, and contributed significantly to the writing and revision of the manuscript. All authors read and approved the final version of the manuscript.

## Data Availability

All data produced in the present work are contained in the manuscript.

## ACKNOWLEDGMENTS

We are grateful to the families participating in the MSSNG, SSC and TOPMed cohorts. We extend our gratitude to David Yang and Marliette Rodriguez-Matos for their invaluable support in resolving technical issues with coding scripts.

## SUPPLEMENTARY INFORMATION

**Supplementary Tables**

**Supplementary Table 1**

The 15 autism gene lists used in this study.

**Supplementary Table 2**

The CWAS categories that showed differences in a comparison of neurotypical compared with neurodivergent individuals.

**Supplementary Table 3**

The CWAS categories that showed differences in a comparison of low (≤70) compared with typical IQ (>70) individuals.

**Supplementary Table 4**

Annotation of clinical and *de novo* regulatory variant (DNRV) properties of the individuals tested in this study.

**Supplementary Table 5**

Annotation of the properties of the DNRVs linked to the individuals tested in this study.

## Supplementary Figures

**Supplementary Figure 1:**
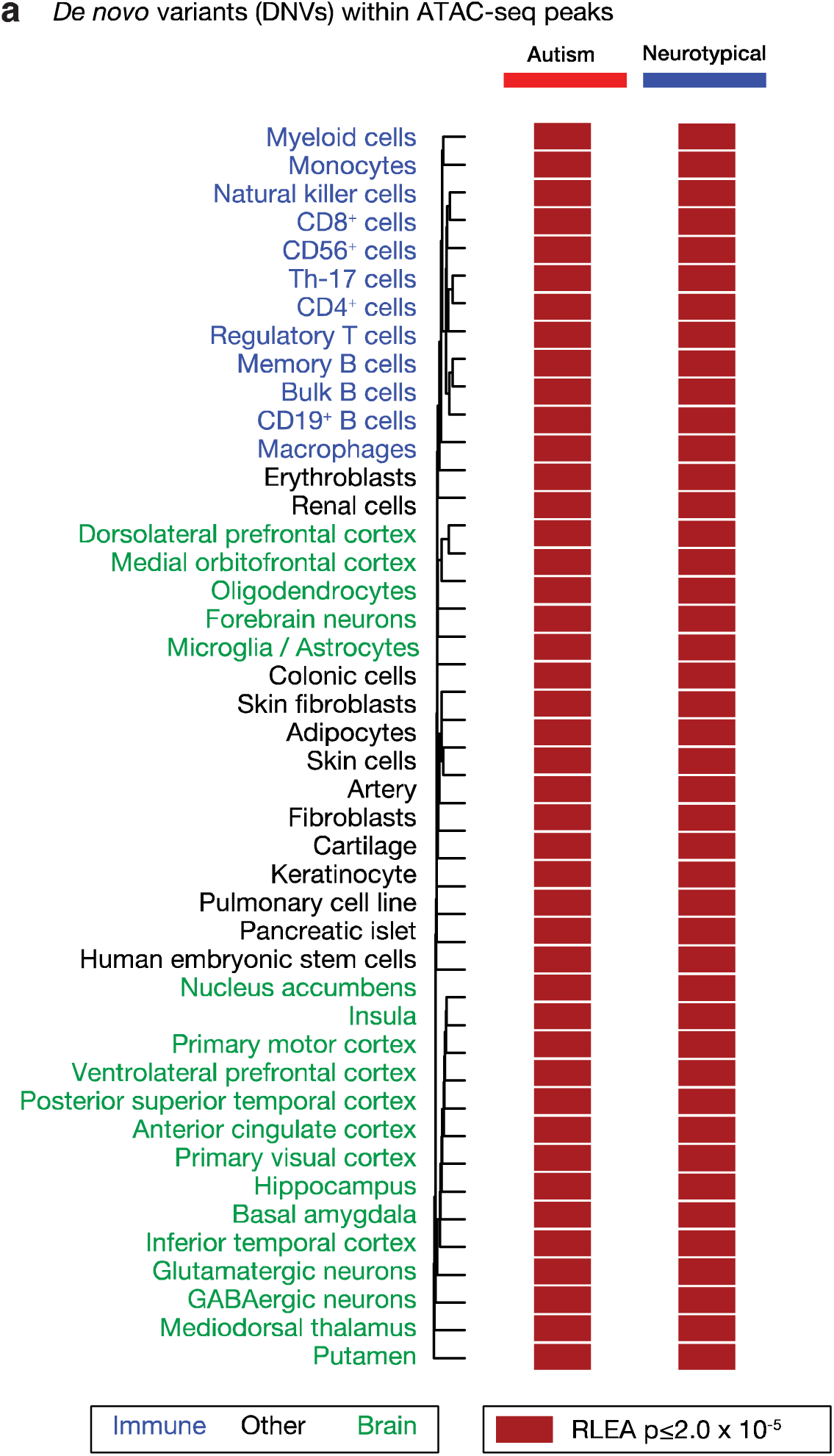
Regulatory landscape enrichment analysis (RLEA) demonstrates global DNV enrichment across the *cis*-regulatory regions of diverse cell types. The RLEA approach was used to test for non-random over-representation of DNVs in open chromatin regions of 44 cell types. Every cell type revealed significant enrichment for DNVs in their open chromatin regions, independent of neurodevelopmental status (threshold *p* ≤ 2.0 x 10^-5^).

**Supplementary Figure 2:**
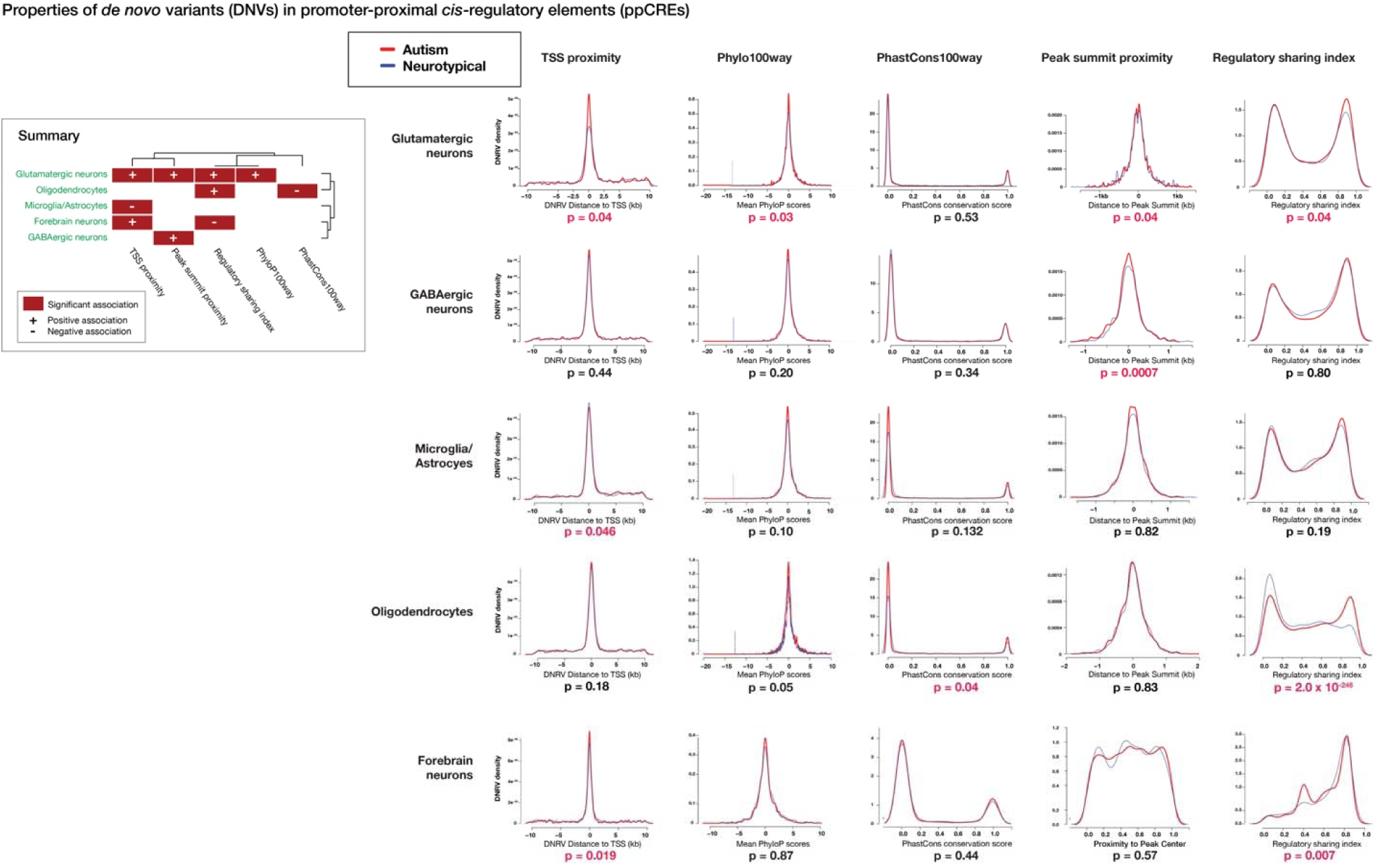
Comparison between the autism and neurotypical groups demonstrates that the properties of glutamatergic regulatory loci are the most distinguishing between groups. The TSS proximity, peak summit proximity, conservation status (PhyloP and PhastCons100way) and regulatory sharing index from regulatory loci in microglia/astrocytes, oligodendrocytes, glutamatergic, GABAergic, and forebrain neurons were compared between the autism (red) and neurotypical group (blue). Glutamatergic neuron genomic properties were significantly different between neurotypical and individuals with autism in all categories except PhastCons100way. Statistical significance was determined using a two-tailed Student’s (p <0.05). A Wilcoxon-Mann-Whitney test was used to assess significance in data with bimodal distributions (PhastCons and regulatory sharing index).

**Supplementary Figure 3:**
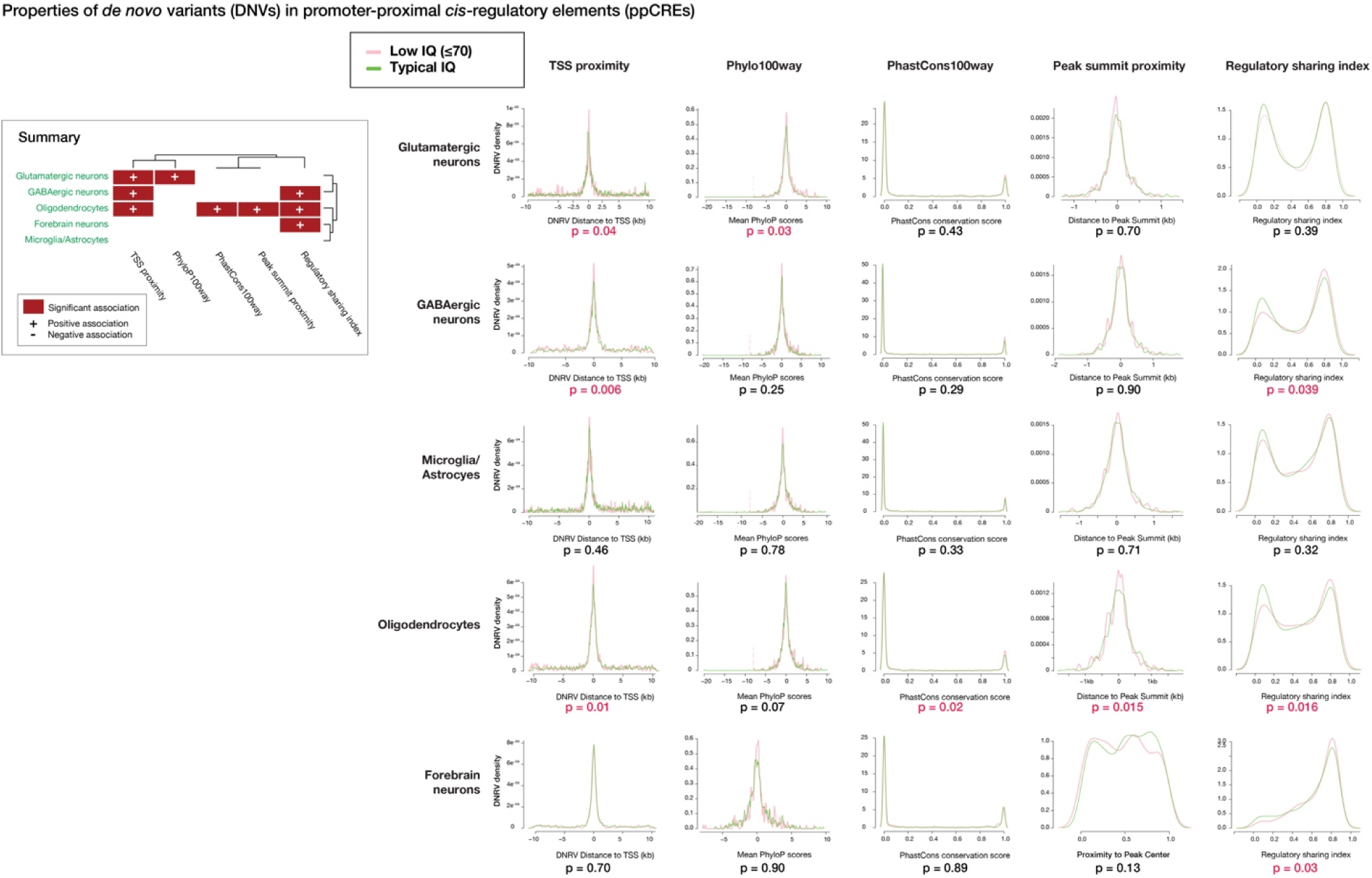
Comparison between low and typical IQ reveals significant differences in the genomic properties of brain cell types. The TSS proximity, peak summit proximity, conservation status (PhyloP and PhastCons100way) and regulatory sharing index from regulatory loci in microglia/astrocytes, oligodendrocytes, glutamatergic, GABAergic, and forebrain neurons were compared between low (pink) and typical IQ autism (green). Oligodendrocyte genomic properties were significantly different between both autism groups in all descriptive properties except PhyloP conservation score (*p*: 0.07). Statistical significance was determined using a two-tailed Student’s (p <0.05). A Wilcoxon-Mann-Whitney test was used to assess significance in data with bimodal distributions (PhastCons and regulatory sharing index).

**Supplementary Figure 4:**
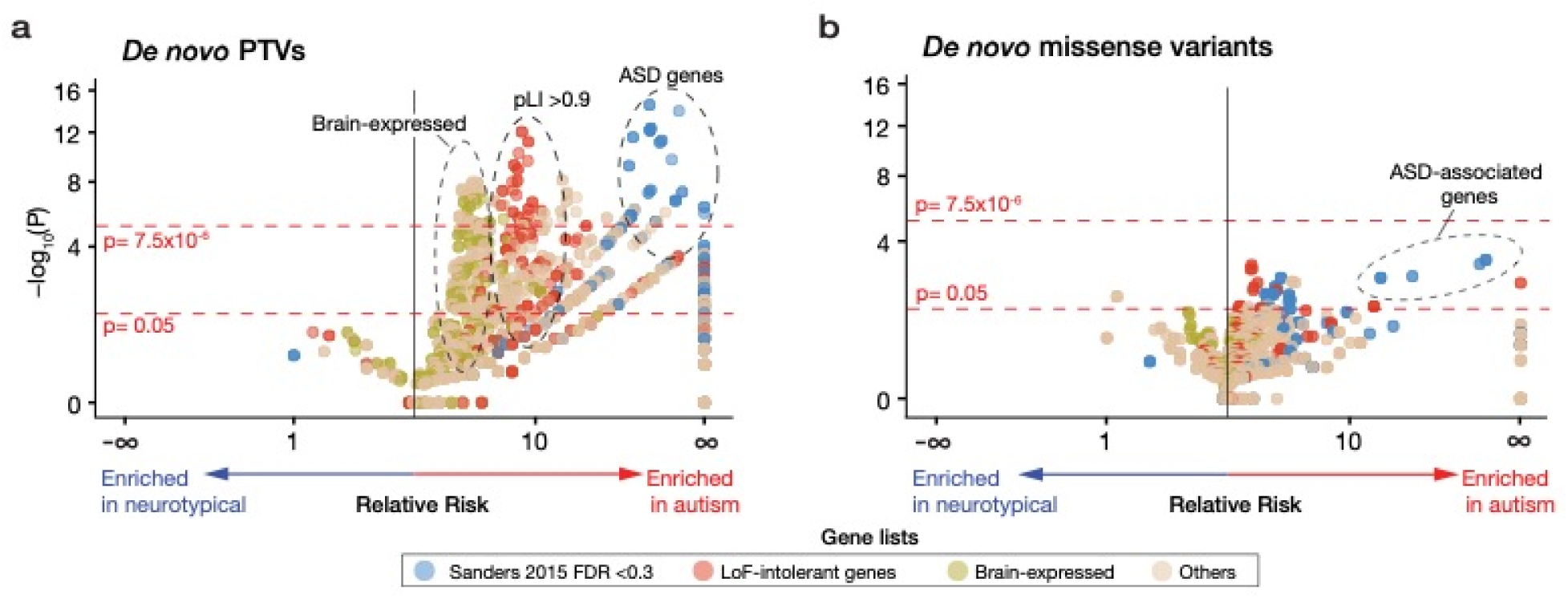
Category-wide association study of the SSC cohort replicates the contributions of *de novo* protein truncating and missense variants in autism. (a) CWAS was performed to replicate the previous findings of An *et al.,* 2019 using 1,902 families from the SSC cohort. *De novo* protein truncating variants (PTVs) are enriched across brain expressed, autism, and evolutionarily conserved (pLI >0.9) genes in individuals with autism (right side). The horizonal dashed red lines indicate the nominal significance threshold (*p* <0.05) and after correcting for the number of tests (*p* < 7.5 x 10^-6^). PTVs were most enriched near “ASD genes” which are represented in the Sanders risk genes TADA (FDR <0.3) gene list. **(b)** The equivalent CWAS is shown for *de novo* missense variants. Although *de novo* missense variants were similarly enriched in individuals with autism (*p* <0.05), they did not surpass the significance threshold after correcting for multiple testing.

**Supplementary Figure 5:**
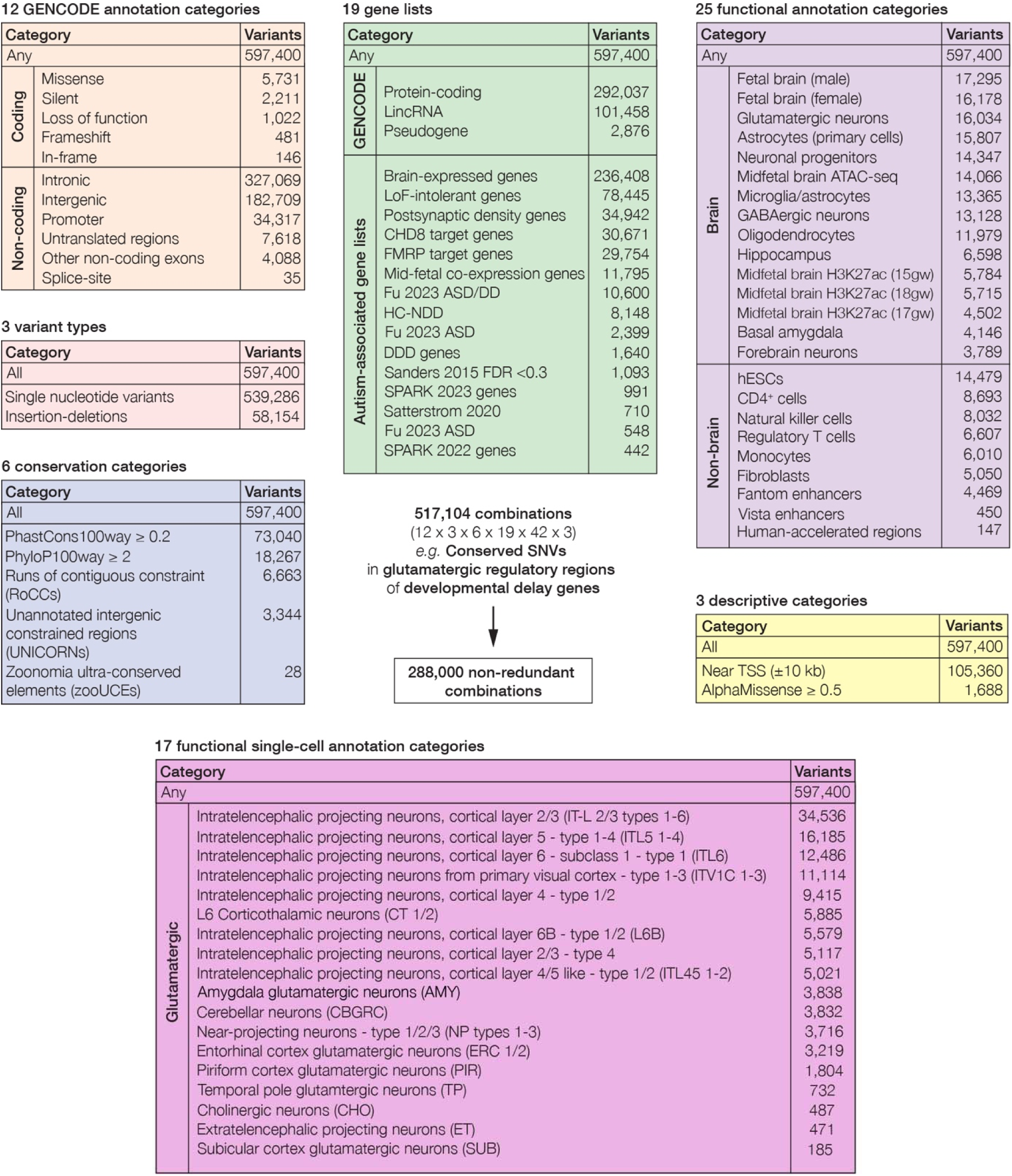
Summarization of annotation categories used to analyse 597,400 variants by CWAS. Seven groups of annotations were defined composed of 25 functional annotations from cell/tissue types (purple) or pseudobulked single nuclear ATAC-seq from 18 cell subtypes (dark pink), 19 gene lists (green), 12 GENCODE annotations (orange), 6 conservation measures (blue), 3 descriptive parameters (yellow), 3 variant types (light pink). These variables provided 517,104 combinations, of which 288,000 were unique.

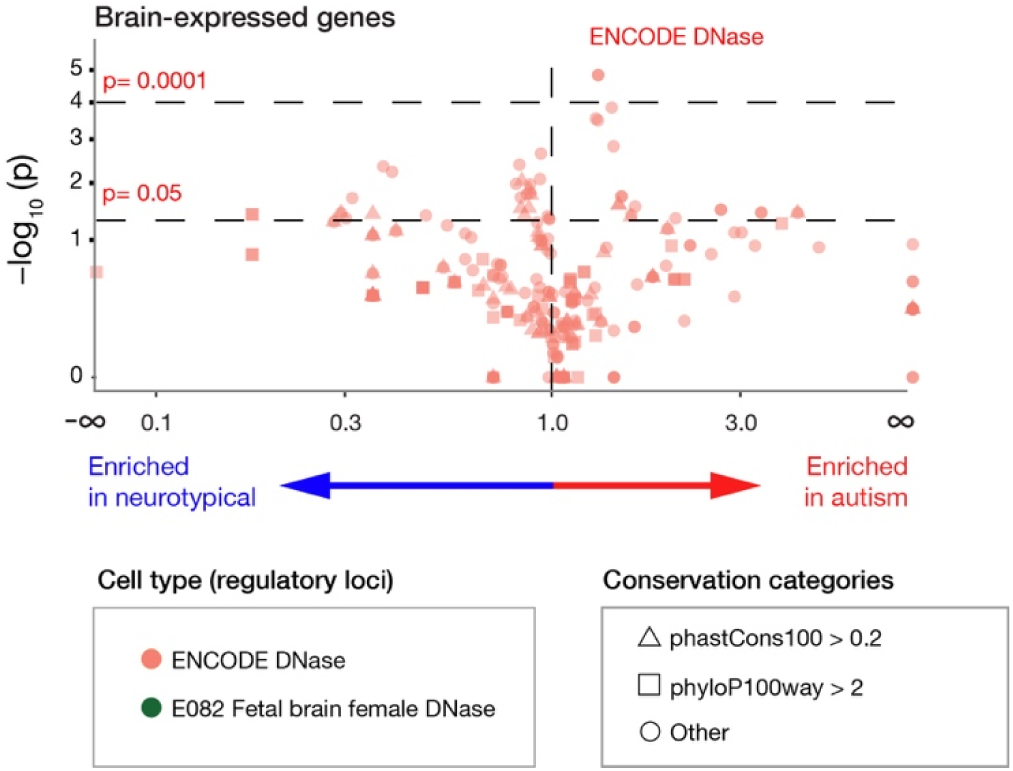
CWAS reveals DNV enrichment at CREs of glutamatergic neurons near Brain-expressed genes in individuals with autism. DNVs in ppCREs defined by ENCODE DNase hypersensitivity data (red points) near known brain-expressed genes are significantly enriched in the autism group.

